# Hospitalization, death, and probable reinfection in Peruvian healthcare workers infected with SARS-CoV-2: a national retrospective cohort study

**DOI:** 10.1101/2022.06.06.22276070

**Authors:** Willy Ramos, Nadia Guerrero, Omar Napanga Saldaña, José Medina, Manuel Loayza, Jhony A. De La Cruz-Vargas, María Vargas, Luis Ordóñez, Yovana Seclén, Carlos Álvarez-Antonio, Juan Arrasco

**Affiliations:** Centro Nacional de Epidemiología, Prevención y Control de Enfermedades. Lima, Perú; Instituto de Investigaciones en Ciencias Biomédicas (INICIB). Facultad de Medicina, Universidad Ricardo Palma. Lima, Perú; Unidad de Post Grado. Facultad de Medicina, Universidad Nacional Mayor de San Marcos. Lima, Perú; Centro de Prevención y Control de Enfermedades, Dirección Regional de Salud de Loreto. Iquitos, Perú

**Keywords:** Healthcare workers, COVID-19, death, reinfection

## Abstract

**OBJECTIVE:** To determine if occupation is a risk factor for probable reinfection, hospitalization, and death from COVID-19 in Peruvian healthcare workers infected with SARS-CoV-2.

**MATERIAL AND METHODS:** Retrospective cohort study. Healthcare workers who presented SARS-CoV-2 infection between March 1, 2020 and August 9, 2021 were included. Occupational cohorts were reconstructed from the following sources of information: the National Epidemiological Surveillance System, molecular tests (NETLAB), results of serology and antigen tests (SICOVID-19), National Registry of Health Personnel (INFORHUS) and National Information System of Deaths (SINADEF). The incidence of probable reinfection, hospitalization, and death from COVID-19 was obtained in the cohorts of health auxiliaries and technicians, nursing staff, obstetricians, physicians, and other healthcare workers. We evaluated whether occupation was a risk factor for probable reinfection, hospitalization, and death from COVID-19 using a log-binomial generalized linear model, obtaining the adjusted relative risk (RR _AJ_).

**RESULTS:** 90,672 healthcare workers were included. 8.1% required hospitalization, 1.7% died from COVID-19, and 2.0% had probable reinfection. A similar incidence of probable reinfection was found in the 5 cohorts (1.9%-2.2%). Physicians had a higher incidence of hospitalization (13.2%) and death (2.6%); however, they were also those who presented greater susceptibility linked to non-occupational variables such as age and comorbidities. The multivariate analysis found that physicians (RR=1.691; CI 95: 1.556–1.837) had a higher risk of hospitalization and that the occupation of health technician and assistant was the only one that constituted a risk factor for mortality from COVID-19 (RR =1.240; 95% CI: 1.052–1.463).

**CONCLUSIONS:** Peruvian health technicians and auxiliaries have a higher risk of death from COVID-19 linked to their occupation, while doctors have higher mortality due to non-occupational factors. Physicians had a higher risk of hospitalization independent of the presence of comorbidities and age; likewise, all occupations had a similar risk of probable reinfection.

## INTRODUCTION

Since the beginning of the COVID-19 pandemic, healthcare workers have been identified as one of the groups at greatest risk due to their direct exposure to SARS CoV-2 as part of health care ^1-6;^ Thus, the first reports made in China and later in European countries ^(7-11)^ evidenced the rapid and exponential transmission in healthcare personnel. As of May 7, 2020, health personnel accounted for 22% of COVID-19 cases in Spain ^9^. In the Region of the Americas, according to the Pan American Health Organization, as of August 20, 2021, 1,792,212 cases and 10,302 deaths from COVID-19 were reported in healthcare workers ^6^.

The increased risk of SARS-CoV-2 infection in healthcare workers is explained by their greater exposure to patients with COVID-19, to the procedures that generate aerosol, to the prolonged use of personal protective equipment or to the lack of any of them (masks, gloves, gown, eye protection) ^12-13^. For this reason, the World Health Organization, as part of their public health strategies to safeguard their health and guarantee the continuity of health care, prioritized the vaccination of healthcare workers ^14^.

Although it is true that healthcare workers have a significantly higher risk than the general population for SARS-CoV-2 infection as well as for adverse outcomes, this may vary according to occupational groups and according to the outcome that is evaluated (reinfection, hospitalization, and death). Likewise, the risk granted by health care is added to that derived from individual risk factors such as age, sex, and diagnosis of comorbidities, among others. ^2,4,5,7^

Peru is one of the countries with the most adverse results in the control of the SARS-CoV-2 pandemic, having reached the highest cumulative global mortality rate per hundred thousand inhabitants since the first wave, a phenomenon that continues to this day^15^. It is to be assumed that this has also been transferred to healthcare workers, during the time of the greatest pandemic activity in which the demand for cases far exceeded the supply of health services, in which there was a significant deficit of human resources, personal protective equipment and equipment which would lead to the collapse of health services^16-18^. To reduce the transmission of SARS-CoV-2, the Peruvian government established various measures such as remote work for tasks that could be carried out from home and for people belonging to risk groups, which included healthcare workers ^19^.

For this reason, it is necessary to assess the risk of adverse outcomes of SARS-CoV-2 infection in Peruvian healthcare workers, discriminating between the risk conferred by occupation and the risk attributable to non-occupational risk factors. The objective of the present investigation was to determine whether occupation and other non-occupational variables constituted risk factors for probable reinfection, hospitalization, and death from COVID-19 in cohorts of Peruvian healthcare workers infected with SARS-CoV-2.

## MATERIAL AND METHODS

National retrospective cohort study. The studied population consisted of healthcare workers who presented SARS-CoV-2 infection between March 1, 2020 and August 6, 2021. All healthcare workers who presented SARS-CoV-2 infection were included in the study, whether they be symptomatic or asymptomatic, as long as they were positive or reactive to RT-PCR, antigen, or serology testing. Healthcare workers older than 70 years and those with inconsistent records were excluded from the study. Sampling was not carried out, we worked with all healthcare workers who met the inclusion and exclusion criteria because they were accessible through secondary sources.

The cohort of healthcare workers infected with SARS-CoV-2 was reconstructed from the following sources of information: National Epidemiological Surveillance System (NOTI COVID-19), molecular test results (NETLAB), serology results, and antigen (SICOVID-19), National Registry of Health Personnel (INFORHUS) and National Deaths Information System (SINADEF), uniting them to form a database.

From the resulting database, the incidence of probable reinfection by SARS-CoV-2 as well as hospitalization and death by COVID-19 was obtained in five occupational cohorts: health assistants and technicians, nursing staff, obstetricians, doctors, and others healthcare workers. This last category grouped together biologists, health career interns, veterinarians, nutritionists, dentists, chemical-pharmacists, medical technologists, and psychologists. It was evaluated if this occupation constituted a risk factor for probable reinfection by SARS-CoV-2, hospitalization, and death by COVID-19. The following occupational and non-occupational variables were also evaluated:

- Probable SARS-CoV-2 reinfection: Work institution, region where hospital is located, direct contact with COVID-19 cases in the work environment, age, sex, and diagnosis of comorbidities.
- Hospitalization for COVID-19: Work institution, age, diagnosis of comorbidities, within the work institution, the hospitals of the Ministry of Health and regional governments (MINSA/GORE), Social Security of Peru (EsSalud), National Police and Armed Forces (PNP/FF. AA) and private establishments were considered.
- Death by COVID-19: Area of work, years working there, age, sex, and diagnosis of comorbidities.

A health worker was considered to have probable reinfection if he or she presented more than one positive or reactive laboratory result separated by at least 3 months ^20^. Death due to COVID-19 was defined as death occurring as a consequence of the natural history or clinical course of the disease (without recovery period) and must meet at least one of the following criteria ^20^:

- Virological: Death in a health worker with clinical disease who dies within 60 days after a molecular (RT-PCR) or reactive antigen test for SARS-CoV-2.
- Serologic: Death in an infected health worker with clinical disease who dies within 60 days of a positive IgM or IgM/IgG serologic test for SARS-CoV-2.
- Radiological: Death in a health worker infected health worker with clinical disease who presents a radiological, tomographic, or nuclear magnetic resonance image compatible with COVID-19 pneumonia.
- Epidemiological link: Death in a health worker with pneumonia that has an epidemiological link with a case of COVID-19.
- Epidemiological investigation: Death in a health worker suspected of COVID-19 that is verified by epidemiological investigation.
- Clinical criteria: Death in a suspected case of COVID-19 with a clinical picture compatible with the disease.
- Death certificate: Death in a health worker who has a death certificate in which the diagnosis of COVID-19 is presented as the cause of death.

Descriptive statistics were performed based on the obtained frequencies, percentages, measures of central tendency and dispersion. Bivariate statistics were performed with Pearson’s Chi square test, which was used to compare proportions. To assess the risk of developing adverse outcomes of SARS-CoV-2 infection (probable reinfection, hospitalization, and death), multivariate statistics were performed with a log-binomial generalized linear model, obtaining the adjusted relative risk (RR _AJ_) for other covariates or potentially confounding variables as well as their confidence intervals. In this model, probable reinfection, hospitalization, and death were considered dependent variables, and occupational and non-occupational variables (older adults, male gender, and presence of comorbidities) were considered independent. The calculations were made with a confidence level of 95%.

In regard to the ethical aspects, the study did not require informed consent because it was based on secondary sources. The confidentiality of the information obtained from the healthcare workers was guaranteed and was used only for the purposes of this study. The research was approved by the Research Ethics Committee of the Ricardo Palma University, School of Medicine (Expedited Review: PI-003-2022).

## RESULTS

### CHARACTERISTICS OF THE HEALTHCARE WORKERS

The study included 90,672 healthcare workers who presented SARS-CoV-2 infection during the study period, with health technicians and auxiliaries as well as nursing staff being the most frequent occupational groups.

Regarding personal characteristics, the highest gender frequency was female and 8.9% had some comorbidity. In the case of occupational characteristics, the highest frequency of healthcare workers labor status was contracted workers, they labored in MINSA/GORE establishments, in the department of Lima and 17.2% reported having direct contact with cases of COVID-19 in their work environment. Likewise, 85.9% presented symptomatic disease, the most used tests for confirmation being serology and RT-PCR. This is shown in table 1.

**TABLE 1:** Non-occupational, occupational, clinical and laboratory characteristics of healthcare workers infected with SARS-CoV-2 in Peru.

| FEATURES | FREQUENCY | % |
| --- | --- | --- |
| NON-OCCUPATIONAL |  |  |
| Sex |  |  |
| Male | 26878 | 29.6 |
| Female | 63794 | 70.4 |
| Age group |  |  |
| 18-29 | 17604 | 19.4 |
| 30-65 | 71147 | 78.5 |
| 65 and over | 1921 | 2.1 |
| Comorbidity |  |  |
| Heart disease | 2456 | 2.7 |
| Obesity | 2156 | 2.4 |
| Bronchial asthma | 1920 | 2.1 |
| Diabetes | 1540 | 1.7 |
| Lung disease | 384 | 0.4 |
| Cancer | 231 | 0.3 |
| Renal disease | 192 | 0.2 |
| Liver disease | 181 | 0.2 |
| Neurological disease | 141 | 0.2 |
| Immunodeficiency | 83 | 0.1 |
| Any comorbidity | 8112 | 8.9 |
| OCCUPATIONAL |  |  |
| Occupation |  |  |
| Technician and health assistant | 34928 | 38.5 |
| Nursing staff | 23262 | 25.7 |
| Midwife | 6276 | 6.9 |
| Physician | 16181 | 17.8 |
| Other health professionals | 10025 | 11.1 |
| Labor Status |  |  |
| Contracted worker | 44599 | 49.2 |
| Temporary worker | 34065 | 37.6 |
| Resident doctor | 1553 | 1.7 |
| Rural Service Doctor | 1993 | 2.2 |
| Intern | 863 | 1.0 |
| Not specified | 7599 | 8.4 |
| Work Institution |  |  |
| MINSA/GORE | 64284 | 70.9 |
| ESSALUD | 16061 | 17.7 |
| PNP / Armed Forces | 3470 | 3.8 |
| Others | 4764 | 5.3 |
| Private Establishments | 2093 |  |
| Work region |  |  |
| Lima | 33712 | 37.2 |
| Rest of the Coast | 19896 | 21.9 |
| Mountain range | 25806 | 28.5 |
| Jungle | 11258 | 12.4 |
| Direct contact with COVID-19 cases in the workplace |  |  |
| Yes | 15612 | 17.2 |
| No | 75060 | 82.0 |
| CLINICS AND LABORATORIES |  |  |
| Clinical Presentation |  |  |
| Symptomatic | 77868 | 85.9 |
| Asymptomatic | 12804 | 14.1 |
| Positive or reactive diagnostic test |  |  |
| RT-PCR | 43354 | 47.8 |
| Antigen test | 13475 | 14.9 |
| Serology | 48351 | 53.3 |

### ADVERSE OUTCOMES OF SARS-CoV-2 INFECTION

When evaluating the adverse results of SARS-CoV-2 infection in healthcare workers, it was observed that 8.1% required hospitalization and 1.7% died from COVID-19; while 2.0% presented probable reinfection. A similar incidence of probable reinfection was observed in the different occupational groups (between 1.9% and 2.2%); while it was the doctors who presented the highest incidence of hospitalization (13.2%) and death (2.6%) (table 2). Physicians were also the ones who presented greater susceptibility not linked to occupational variables. In this sense, 3.5% of doctors were older adults compared to 2.1% of health technicians and assistants, 1.6% of nursing staff, 1.0% of obstetricians and 1.5% of the rest of healthcare workers (Pearson’s Chi-square test; p<0.001). Likewise, 13.1% of doctors had comorbidities compared to 7.8% of health technicians and auxiliaries, 8.6% of nursing staff, 8.1% of obstetricians, and 7.7% of the rest of healthcare workers (Pearson’s Chi-square test; p<0.001).

**TABLE 2:** Adverse outcomes of SARS-CoV-2 infection in healthcare workers in Peru.

| ADVERSE OUTCOME | FREQUENCY | INCIDENCE (%) |
| --- | --- | --- |
| Probable reinfection |  |  |
| Technician and health assist. | 700 | 2.0 |
| Nursing staff | 485 | 2.1 |
| Midwife | 135 | 2.2 |
| Physician | 322 | 2.0 |
| Other health professionals | 189 | 1.9 |
| Total | 1831 | 2.0 |
| Hospitalization |  |  |
| Technician and health assist. | 2595 | 7.4 |
| Nursing staff | 1602 | 6.9 |
| Midwife | 328 | 5.2 |
| Physician | 2130 | 13.2 |
| Other health professionals | 662 | 6.6 |
| Total | 7317 | 8.1 |
| Death |  |  |
| Technician and health assist. | 664 | 1.9 |
| Nursing staff | 224 | 1.0 |
| Midwife | 55 | 0.9 |
| Physician | 418 | 2.6 |
| Other health professionals | 161 | 1.6 |
| Total | 1522 | 1.7 |

The risk of probable reinfection was similar in the different occupational cohorts of healthcare workers. Regardless of occupation, a higher risk of probable reinfection was observed in those who worked in MINSA/GORE establishments, in those who worked outside of Lima, in those who had direct contact with COVID-19 cases in their work environment, and in those who had some comorbidity (Table 3).

**TABLE 3:**
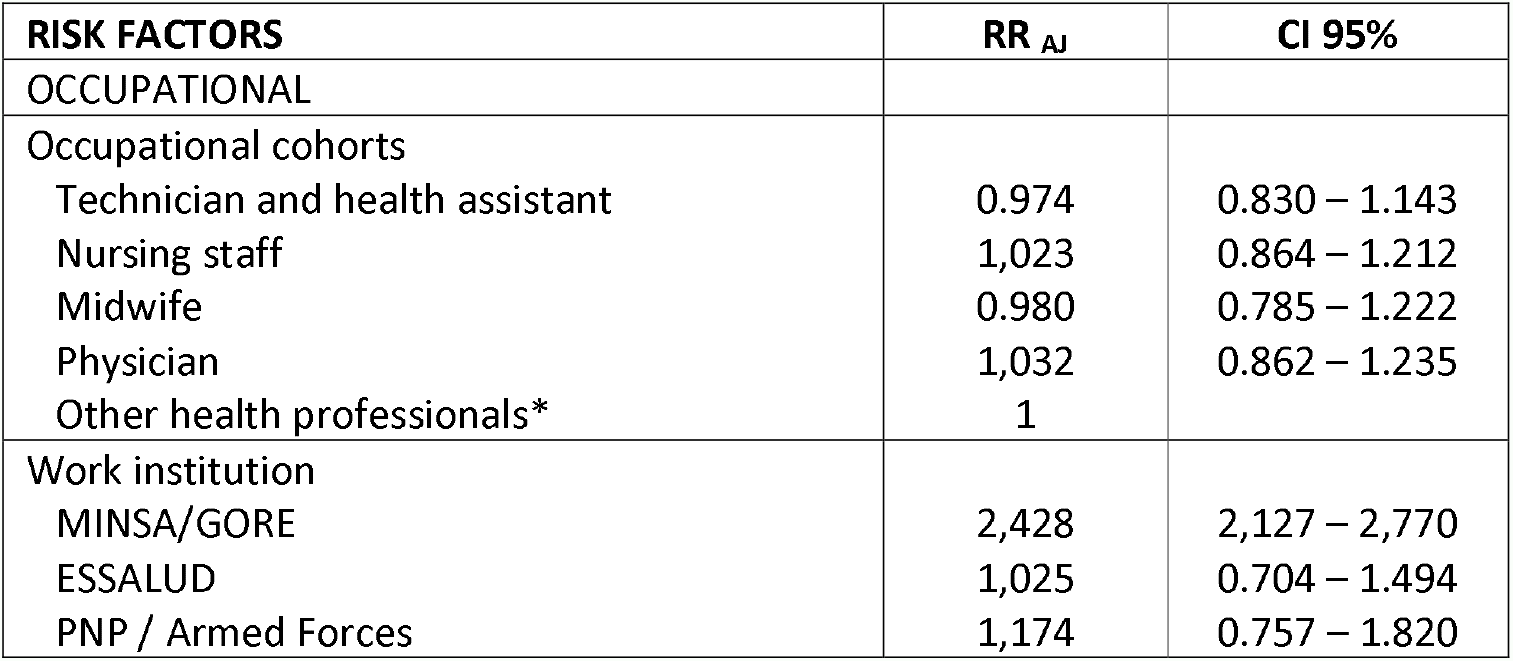
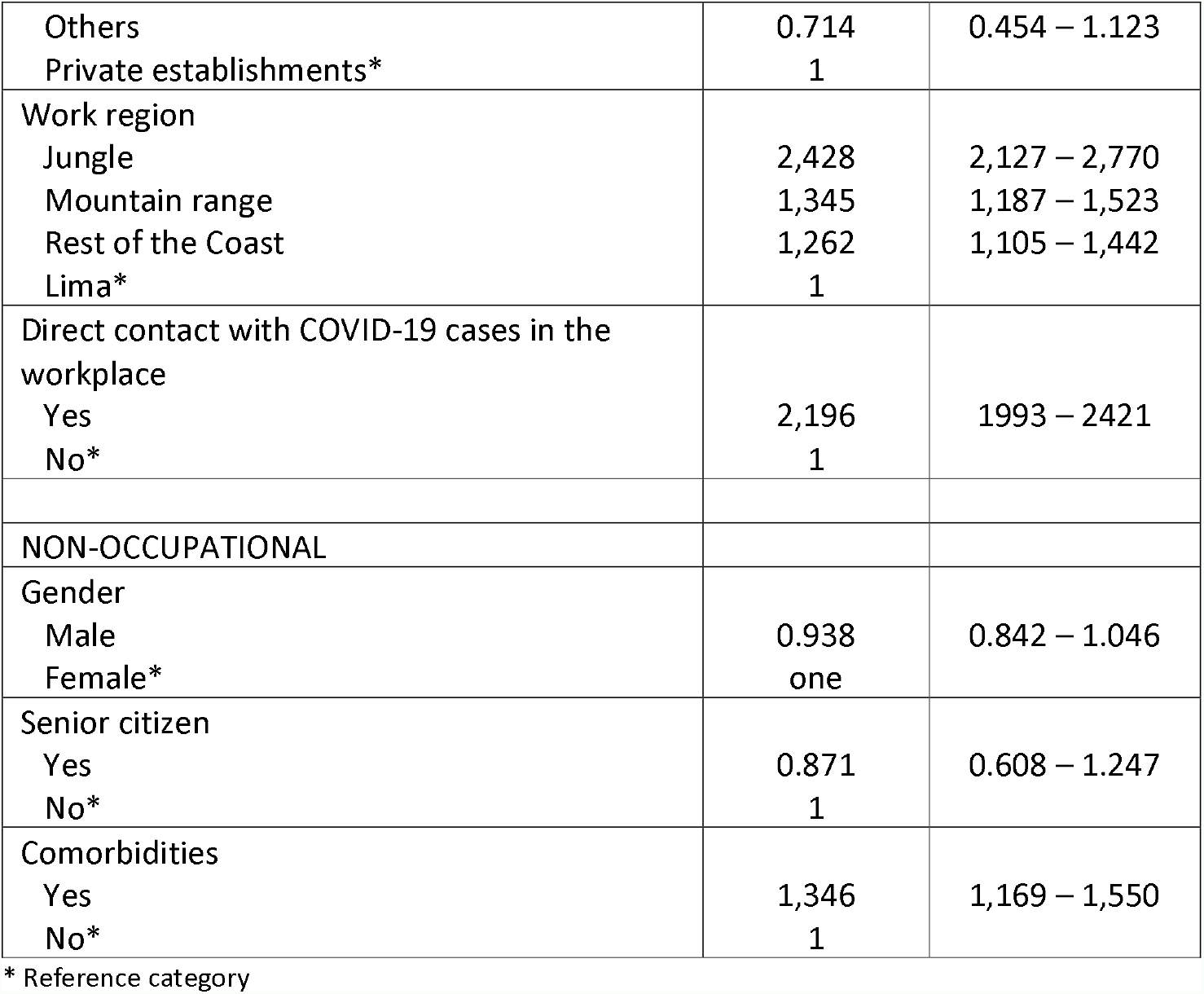
Multivariate analysis of occupation and other possible risk factors for probable SARS-CoV-2 reinfection in healthcare workers.

The multivariate analysis shows that doctors and, to a lesser extent, health technicians and auxiliaries presented a higher risk of hospitalization. Regardless of the occupation, a higher risk was observed in those who worked in Lima and other parts of the coast, in those who had some comorbidities and in the older adults, the latter being the ones with the highest risk (RR _AJ_ =2.947). This is shown in table 4.

**TABLE 4:** Multivariate analysis of occupation and other possible risk factors for hospitalization for COVID-19 in healthcare workers.

| RISK FACTORS | RR<sub>AJ</sub> | CI 95% |
| --- | --- | --- |
| OCCUPATIONAL |  |  |
| Occupational cohorts |  |  |
| Technician and health assistant | 1,091 | 1,005 – 1,184 |
| Nursing staff | 1,015 | 0.931 – 1.107 |
| Midwife | 0.817 | 0.719 – 0.928 |
| Medical | 1,691 | 1556 – 1837 |
| Other health professionals* | 1 |  |
| Work region |  |  |
| Lima | 1,272 | 1,180 – 1,370 |
| Rest of the Coast | 1,126 | 1,039 – 1,221 |
| Mountain range | 1,001 | 0.923 – 1.085 |
| Jungle* | 1 |  |
| PERSONAL |  |  |
| Older adult |  |  |
| Yes | 2,947 | 2,760 – 3,148 |
| No* | 1 |  |
| Comorbidities |  |  |
| Yes | 2,338 | 2,221 – 2,461 |
| No* | 1 |  |
| Likely reinfection |  |  |
| Yes | 0.969 | 0.830 – 1.130 |
| No* | 1 |  |

\* Reference category

The trend of deaths from COVID-19 in healthcare workers compared to the trend in the general Peruvian population is shown in Figure 1. After controlling for other non-occupational and occupational variables, it was found that the occupation of technician and assistant in health was the only one that constituted a risk factor for mortality from COVID-19 (RR AJ =1.240; 95% CI: 1.052 – 1.463). Likewise, there was a higher risk of death for those who worked in Lima and the rest of the coast, in those who had some comorbidity and in older adults, the latter being the ones with the highest risk (Table 5).

**TABLE 5:** Multivariate analysis of occupation and other possible risk factors for death from COVID-19 in healthcare workers.

| RISK FACTORS | $RR_{AJ}$ | CI 95% |
| --- | --- | --- |
| OCCUPATIONAL |  |  |
| Occupational cohorts |  |  |
| Technician and health assistant | 1,240 | 1,052 – 1,463 |
| Nursing staff | 0.840 | 0.687 – 1.028 |
| Midwife | 0.857 | 0.632 – 0.161 |
| Physician | 0.925 | 0.776 – 1.102 |
| Other health professionals* | 1 |  |
| Work region |  |  |
| Jungle | 1,442 | 1,229 – 1,693 |
| Mountain range | 0.982 | 0.822 – 1.174 |
| Rest of the Coast | 1,173 | 0.982 – 1.401 |
| Lima* | 1 |  |
| Year of employment |  |  |
| 2021 | 1,272 | 1,156 – 1,400 |
| 2020* | 1 |  |
| INDIVIDUALS |  |  |
| Gender |  |  |
| Male | 2,722 | 2,442 – 3,035 |
| Female* | 1 |  |
| Senior Citizen |  |  |
| Yes | 8,959 | 7,996 – 10,037 |
| No* | 1 |  |
| Comorbidity |  |  |
| Yes | 2,523 | 2,271 – 2,824 |
| No* | 1 |  |

\* Reference category

**FIGURE 1:**
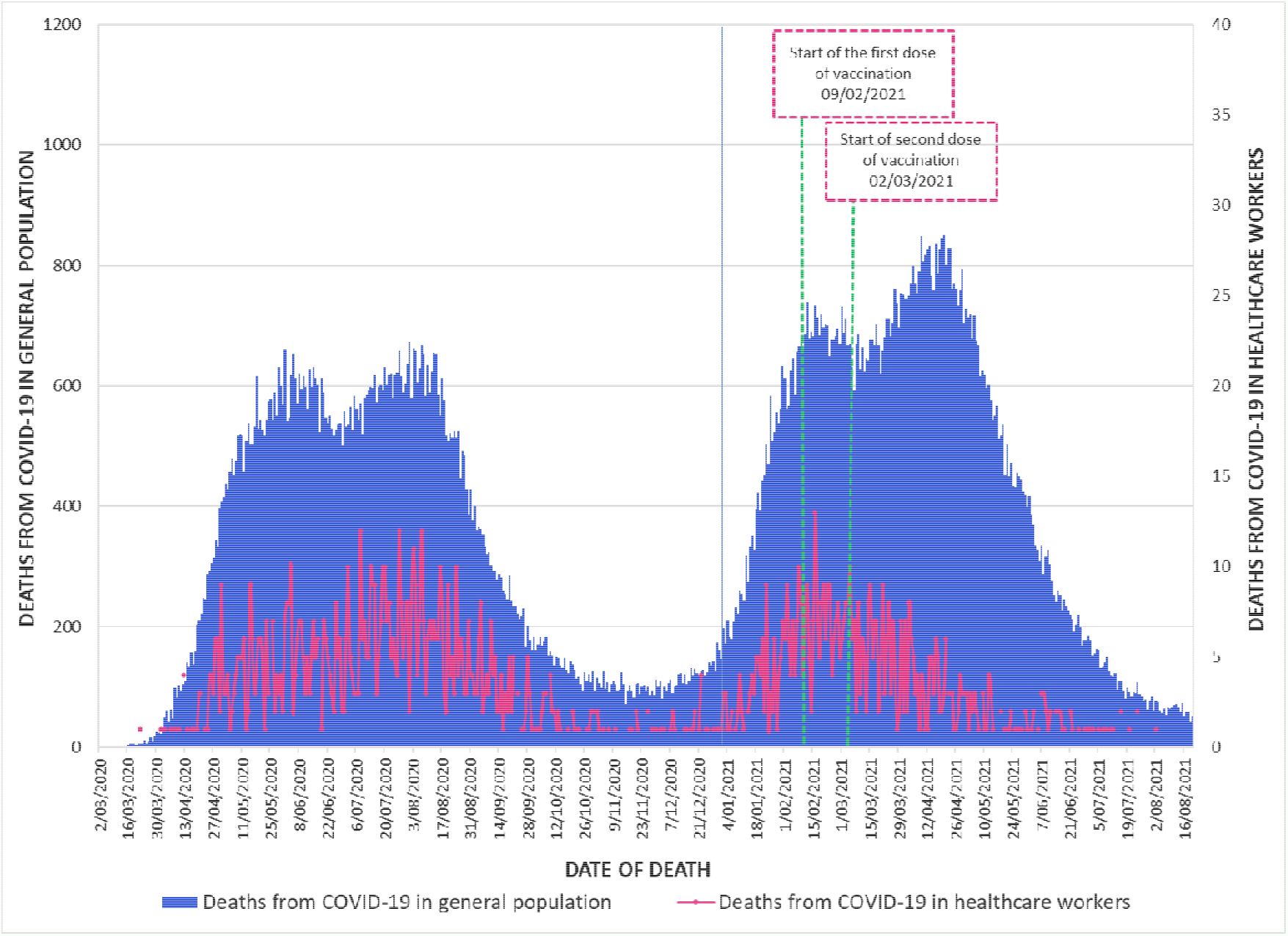
Trend of COVID-19 deaths in healthcare workers compared to the general Peruvian population.

## DISCUSSION

The present investigation shows that cohorts of Peruvian healthcare workers have differences amongst the risk of hospitalization and death from COVID-19 based on their occupation. The strong influence of non-occupational factors that increase the risk of adverse outcomes of SARS-CoV-2 infection is also observed.

Death from COVID-19 represents the main adverse outcome of SARS-CoV-2 infection. The mortality in healthcare workers in Peru infected by SARS-CoV-2 found in our study was 1.7%, which is higher than that reported by Gholami ^21^ who found a mortality of 1.5% in a meta-analysis of 28 studies. A second meta-analysis by Gómez-Ochoa ^22^ that included 97 studies found that mortality from COVID-19 in infected workers was 0.5%, which is notably lower than that reported in this study for Peruvian healthcare workers. It should be noted that the meta-analysis carried out by Gholami included 119,883 healthcare workers infected with SARS-CoV-2 and that carried out by Gómez-Ochoa included 96,813; whereas the Peruvian health worker cohort alone included 90,672 infected healthcare workers. This shows in absolute terms the great affectation of Peruvian healthcare workers by the pandemic, not only due to the high workload, continuous exposure, and lack of personal protective equipment (mainly at the beginning of the pandemic); but also due to the informal nature of work and the worsening labor conditions observed in various countries since before the pandemic, particularly those with low and medium income ^23-26^.

The results of our research show that the cohort with the highest risk of death from COVID-19 derived from their occupation was that of health technicians and auxiliaries, who had a 24% higher risk of dying than other healthcare workers. This is explained because the cohort of health technicians and auxiliaries includes technicians and auxiliaries in nursing, laboratory, dental, pharmacy, nutrition, radiology, rehabilitation, and physical therapy. This occupational cohort, particularly nursing technicians, who collaborate with patient care in consulting rooms, emergencies, hospitalization (including feeding, cleaning, mobility, patient oxygen administration, among others) as well as radiology and laboratory technicians, have close contact with patients or with biological samples^27^. This leads to a higher viral load exposure and would explain their greater overall risk ^28-30^. The bibliographic review does not find studies with the category of health technicians and auxiliaries like that defined in Peru, but an approximation is found in the cohort of Mexican healthcare workers that finds a higher risk of death in medical assistants, laboratory technicians, pharmacy, and radiology staff ^31^.

The case of doctors is particular because they present an occupational risk of dying similar to other healthcare workers; however, they are the ones with the highest mortality (2.6%). This is consistent with the results of a systematic review ^32^ that found that the group with the highest mortality among infected healthcare workers were doctors (6.0%). One possible explanation for this phenomenon is that in Peru, the physician cohort is the one with the highest proportion of older adults and comorbidities compared to the other cohorts of healthcare workers, which could explain their higher mortality from COVID-19, regardless of their occupational risk. Another possible explanation is that physicians have performed diagnostic tests less frequently in the presence of mild disease and more frequently in the presence of moderate and severe disease, which could have biased the results towards greater lethality^33^.

It is observed that non-occupational risk factors lead to a higher risk of death from COVID-19 than occupational factors, the main one is being a older adults; thus, older adults have about 9 times the risk of dying than those under 65 years of age. Male gender, as well as the presence of comorbidities, are risk factors for death from COVID-19, as it occurs in the general population. This agrees with other studies carried out on healthcare workers such as the one carried out by Ferland in 9 European countries^34^ and the one by Robles-Pérez in Mexico^31^. The main comorbidities identified were cardiovascular disease, obesity, bronchial asthma, and diabetes mellitus, representing 8.9% of the total infected. These values are similar to those found in the meta-analysis by Gómez-Ochoa, who found that the prevalence of comorbidities was 7% (95% CI: 4% - 10%).^22^

The trend of deaths from COVID-19 in healthcare workers follows a trend similar to that of the general Peruvian population; however, this correlation breaks down after the start of vaccination. Thus, the trend in deaths decreased after vaccination; while, in the general population, the trend was to increase until reaching the highest value of the pandemic during the second wave. This confirms the results of Escobar-Agreda ^35^ who found a higher survival rate in Peruvian healthcare workers in 2021 after the start of vaccination. This would show the effectiveness of vaccination since without it, the number of deaths from COVID-19 would have increased notably in a similar way to what happened with the general Peruvian population.

In the case of hospitalizations for COVID-19, it was the doctors who showed 69.1% more risk than the cohort of other healthcare workers; while technicians and health assistants had a 9.1% higher risk of being hospitalized. The fact that technicians and health assistants are the ones with the highest risk of death but a modest additional risk for hospitalization could show inequity in access to hospitalization, which could also explain the higher risk of deaths from COVID-19 in this cohort. It is possible that the efforts of the professional associations, in obtaining air transport for their members and the coordination for their referrals, as is the case of the Peruvian Medical Association, has contributed to a timely hospitalization of its members, reducing their mortality ^36^. Unfortunately, there is no school, society or association of health technicians and auxiliary support in Peru that would ensure the timely hospitalization of its members, which may have put them at a disadvantage with other occupations that do have professional associations. Although it is true that our bibliographic review has not found studies that show inequities in access to hospitalization services in occupational groups of healthcare workers, this is possible since there is evidence of inequities for hospitalization in more disadvantaged or invisible groups during the SARS-CoV-2 pandemic ^37,38^.

Probable reinfection was documented in 1.7% of healthcare workers. It was observed that the risk of probable reinfection was similar in the cohorts of healthcare workers studied; however, other occupational factors were relevant. The greatest risk of probable reinfection was found in those who worked in public establishments of the MINSA/GORE, PNP/FFAA, as well as in those who worked outside the capital, particularly in establishments in the Amazon and the Andean region; a higher risk was also documented in those who had direct contact with COVID-19 cases in their workplace. It is possible that the greater limitations existing in the establishments of the MINSA/GORE, PNP/FAA and outside the capital of Peru, have contributed to the reinfection of healthcare workers during the pandemic’s greatest activity, moments in which there have been documented deficit of personal protective equipment, as well as a greater exposure to COVID-19 due to the overwhelming patient demand^17,35,39-41^.

Within the limitations, it should be stated that our study was carried out using secondary sources, so it is possible that there are some degree of quality problems and underreporting; however, the fact that we considered various sources of information as well as the verification and investigation of deaths would partially compensate for these limitations. Likewise, it was not possible to document which healthcare workers were providing face-to-face care and which were performing remote work. Finally, it was not possible to confirm the reinfection of healthcare workers because in Peru the identification of SARS-CoV-2 lineages is not routinely performed in all cases of infection, so probable reinfection was evaluated. Despite these limitations, we consider that the results obtained are consistent and are close to those observed in Peruvian healthcare workers during the pandemic.

In conclusion, Peruvian health technicians and auxiliaries have a higher risk of death from COVID-19 linked to their occupation; while physicians have a higher risk of death linked to the high frequency of non-occupational risk factors such as age and the diagnosis of comorbidities. Physicians had a higher risk of hospitalization independent of the presence of comorbidities and age; likewise, all occupations had a similar risk of probable reinfection.

## Data Availability

All data produced in the present study are available upon reasonable request to the authors

